# Lack of Causal Effects or Genetic Correlation between Restless Legs Syndrome and Parkinson’s Disease

**DOI:** 10.1101/2021.02.16.21251687

**Authors:** Mehrdad A Estiar, Konstantin Senkevich, Eric Yu, Parizad Varghaei, Lynne Krohn, Sara Bandres-Ciga, Alastair J Noyce, Guy A Rouleau, Ziv Gan-Or

## Abstract

**Background:** Epidemiological studies have reported association between Parkinson’s disease (PD) and restless legs syndrome (RLS).

**Objectives:** We aimed to use genetic data to study whether these two disorders are causally linked or share genetic architecture.

**Methods:** We performed two-sample Mendelian randomization (MR) and linkage disequilibrium score regression (LDSC) using summary statistics from recent genome-wide meta-analyses of PD and RLS.

**Results:** We found no evidence for a causal relationship between RLS (as the exposure) and PD (as the outcome, inverse variance-weighted; b=-0.003, se=0.031, *p*=0.916, F-statistic=217.5). Reverse MR also did not demonstrate any causal effect of PD on RLS (inverse variance-weighted; b=-0.012, se=0.023, *p*=0.592, F-statistic=191.7). LDSC analysis demonstrated lack of genetic correlation between RLS and PD (rg=-0.028, se=0.042, *p*=0.507).

**Conclusions:** There was no evidence for a causal relationship or genetic correlation between RLS and PD. The associations observed in epidemiological studies could be, in part, attributed to confounding or non-genetic determinants.

## Introduction

Restless legs syndrome (RLS) and Parkinson’s disease (PD) are common neurological disorders with a prevalence of 1.9-4.6% and 0.1-2.9% in Europeans, respectively.^1,2^ Epidemiological studies suggest that RLS is more common than expected in PD patients, and PD affects RLS patients more frequently than matched controls or the general population.^3^ Some studies suggest that RLS may be an early clinical manifestation of PD,^4-6^ whereas other studies found no association between RLS and PD.^3^ A recent meta-analyses showed a higher odds for RLS in PD patients compared to controls.^3^ In this study, the previous contradictory results were explained by different inclusion and diagnostic criteria and differences in sex distribution.^3^ However, there are major differences between RLS and PD including clinical, ultrasonographic, functional and neuroimaging aspects, which do not support an association between RLS and PD.^7-10^

Therefore, the true nature of the association between RLS and PD remains unclear. Mendelian Randomization (MR) may help mitigate some of the bias introduced by reverse causation and confounding in traditional observational studies.^11^ In addition, genetic correlation using linkage disequilibrium (LD) score regression (LDSC) may help determine whether different traits have overlapping genetic background, which may explain some of the observed associations between traits.

Here, we used bidirectional MR and LDSC to seek evidence for a causal relationship and/or shared genetic architecture between RLS and PD.

## Methods

### Study population and genetic data

To perform MR and LDSC, we used summary statistics from two recent genome-wide association study (GWAS) meta-analyses of RLS and PD.^12,13^ The RLS summary statistics included data from 15,126 patients and 95,725 controls,^12^ and the PD summary statistics included data from 33,674 cases (15,056 PD patients and 18,618 proxy-cases), and 449,056 controls.^13^ A subset of data (23andMe data) was not included in the PD summary statistics to avoid potential overlap with the RLS data which included 23andMe data. 23andMe participants provided informed consent and participated in the research under a protocol approved by the external AAHRPP-accreted IRB, Ethical & Independent Review Services (E&I Review). The full GWAS summary statistics for the 23andMe discovery data set will be made available through 23andMe to qualified researchers under an agreement with 23andMe that protects the privacy of the 23andMe participants. Please visit https://research.23andme.com/collaborate/#dataset-access/ for more information and to apply to access the data. Information on recruitment procedures and diagnostic criteria is detailed in the original publications.^12,13^ All cases and controls in this study were of European ancestry.

### Power calculation

Power was calculated for detecting an effect size of odds ratio of 1.2 on RLS and PD risk, using online sample size and power calculator for Mendelian randomization with a binary outcome (https://sb452.shinyapps.io/power/).^14^ For all analyses the power was estimated at >80%.

### Mendelian randomization

We performed bidirectional MR, i.e. examining whether RLS is a causal risk factor (exposure) for PD (outcome) and if PD is a causal risk factor for RLS. For each MR analysis, we constructed multi-variant instruments from the independent (“index”) GWAS significant SNPs (p<=5e-08) from the exposure GWAS. In brief, index SNPs were obtained by clumping all GWAS significant SNPs within each LD block using an R^2^ threshold of 0.001 or a distance of 10,000 kb from the index SNP. This process increased the independence of each index SNP based on the above parameters. Additional details regarding the instrument construction and the code used for the analysis are available at https://github.com/gan-orlab/MR_LDSC_RLS-PD.

To calculate the proportion of variability in the exposure explained by the SNPs and to test the strength of the instrument variables (IVs), we used the statistical power for MR analyses (the coefficient of determination, R^2^) and F-statistics tests, as previously described.^15^ In order to perform MR, an estimate of the individual effect of SNPs on the exposure and outcome (RLS and PD, interchangeably) was used to calculate the Wald ratio. Then, the effect estimates were combined using the Inverse-variance weighted (IVW) method, which is a weighted mean of the Wald ratio estimates obtained from each individual SNP separately.^16^

### Sensitivity analyses

To explore whether IVW results might be biased due to violations of MR assumptions and to evaluate the robustness of the results, we used weighted median and MR Egger^16^ estimators as sensitivity analyses. The weighted median estimate provides a reliable pooled estimate assuming that at least half of the weight of the SNPs in the instrument are valid. MR Egger assesses directional pleiotropy similarly to the IVW approach except that the regression slope y-intercept is not constrained to pass through the origin. For each approach, we constructed funnel plots to detect outliers. We evaluated the heterogeneity statistics Q for IVW and Q′ for MR-Egger.

Mendelian Randomization Pleiotropy RESidual Sum and Outlier (MR-PRESSO)^17^ was used to examine outlier SNPs which might occur in the presence of horizontal pleiotropy and correct pooled estimates. Steiger filtering was used to discard SNPs that explain more variance in the outcome than in the exposure. To find all the SNPs that are in LD with the index SNP, the LDmatrix module on the LDlink web tool was used.^18^

### Genetic correlation analyses

To assess the genetic correlation between RLS and PD, we performed LDSC after computing z-scores and formatting data from the two GWASs as previously described.^19,20^ In brief, LDSC calculates genetic correlation between two traits by incorporating LD scores (the more variants in LD with a SNP, the higher the LD score) and GWAS summary statistics (z scores) in a regression model.

## Results

In total, 20 and 55 index SNPs for RLS and PD, respectively, were initially used as IVs for exposure. These IVs explain 3.5% and 2.1% of the risk in RLS and PD. All SNPs were strong instruments for MR analysis as measured by F-statistics (RLS F-statistics=217.5; PD F-statistics=191.7). There was no overlap between the genes where the clumped SNPs are located in both meta-analyses (Supplementary Table 1).

We then performed MR analyses to assess the bidirectional causal relationship between RLS and PD. RLS, as the exposure, was not causally associated with PD (IVW; b=-0.051, se=0.037, *p*=0.172). However, the *p* values of IVW-Q and MR Egger-Q′ tests were 0.034 and 0.025, respectively, raising the possibility of pleiotropic SNP(s) in our dataset, which violates MR assumptions. MR-PRESSO^17^ was applied and a pleiotropic index SNP, rs11860769 (*p*=0.02) was identified when RLS was used as exposure. This SNP has an opposite effect on risk of RLS and PD as was previously shown.^21^ After removing the pleiotropic index SNP (rs11860769), 19 index SNPs were used as IVs for RLS, respectively. Again, there was no causal effect of RLS on PD (b=-0.003, se=0.031, *p*=0.916) or of PD on RLS (b=-0.012, se=0.023, *p*=0.592) with 55 IVs, and the results of sensitivity analyses suggested that there were no additional deviations from the MR assumptions (Table 1, Figure 1, Supplementary Figure 1,2).

**Table 1.** MR analysis between RLS and PD.

| Exposure | Outcome | F | R<sup>2</sup> | MR-PRESSO | Inverse variance weighted |  |  |  | MR Egger |  |  |  | Weighted median |  |  |
| --- | --- | --- | --- | --- | --- | --- | --- | --- | --- | --- | --- | --- | --- | --- | --- |
|  |  |  |  |  | b | se | P | Q test p | b | se | P | Q test p | b | se | P |
| RLS | PD | 217.558 | 0.035 | 0.832 | -0.003 | 0.031 | 0.916 | 0.780 | -0.019 | 0.064 | 0.767 | 0.729 | -0.020 | 0.043 | 0.632 |
| PD | RLS | 191.791 | 0.0315 | 0.662 | -0.012 | 0.023 | 0.592 | 0.300 | -0.002 | 0.050 | 0.958 | 0.269 | -0.011 | 0.036 | 0.749 |

RLS, restless legs syndrome; PD, Parkinson's disease; F, 'strength' of the genetic instrumental variable; R<sup>2</sup>, proportion of variance in exposure variable explained by SNPs; MR-PRESSO, Mendelian Randomization Pleiotropy RESidual Sum and Outlier; MR, Mendelian randomization; b, beta; se, standard error; Q, Cochran's Q test

**Figure 1.**
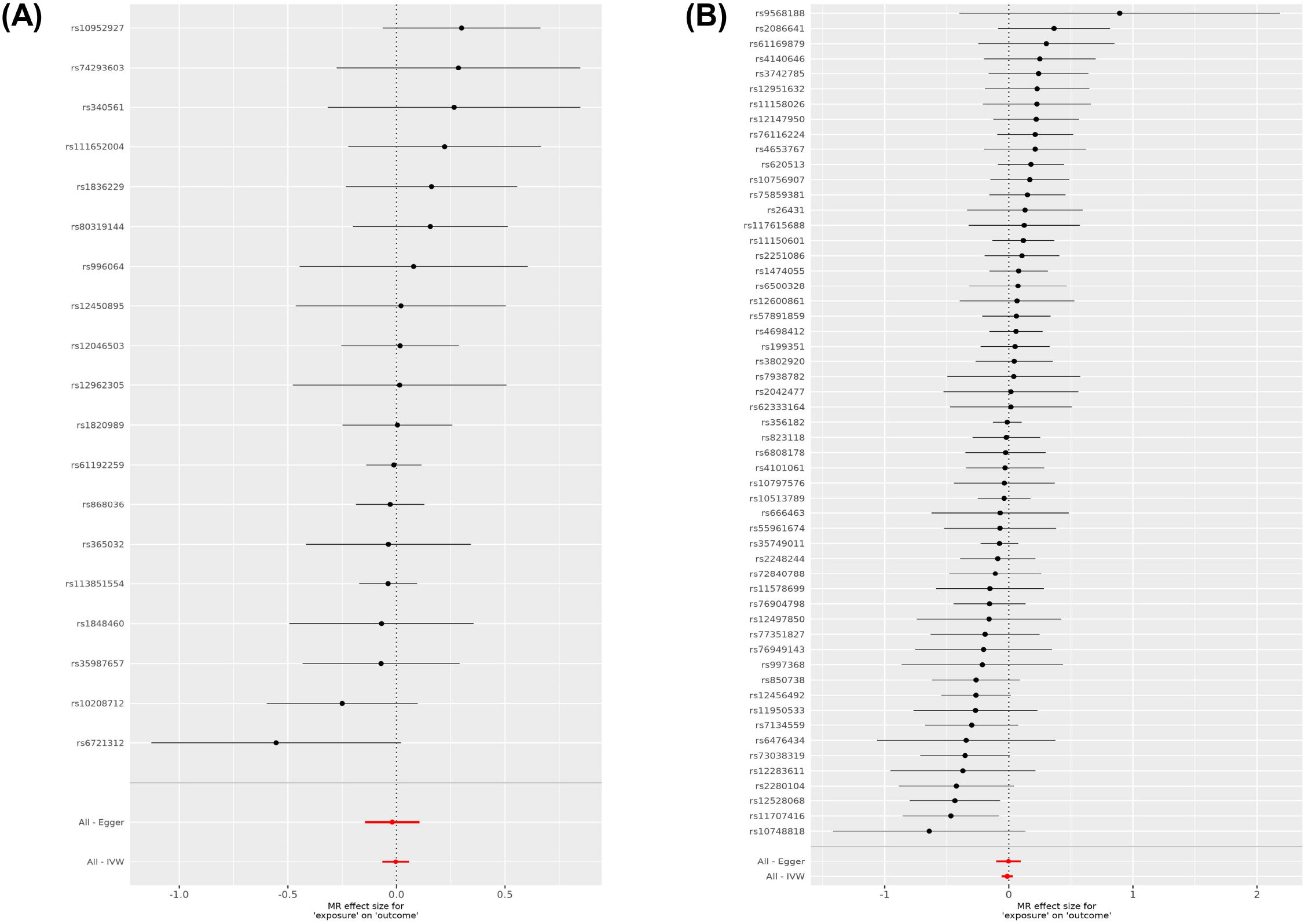
Forest plots showing results from the Mendelian randomization study to evaluate the potential causal relationships between RLS and PD. **A**. Forest plot showing point estimates of RLS as an exposure on PD (outcome). In total, 19 index SNPs were left after excluding the pleiotropic SNPs to construct instrument variables. The black dots represent the causal estimate (b = log odds ratio) of each SNP on the risk of PD. Red dots represent the causal estimate when combining all SNPs together, using MR Egger and IVW methods. Horizontal lines denote 95% CI. **B**. Forest plot showing point estimates of PD as an exposure on RLS (outcome). The instrument variables were constructed by 55 index SNPs. The black dots represent the causal estimate (b = log odds ratio) of each SNP on the risk of RLS. Red dots represent the causal estimate when combining all SNPs together, using MR Egger and IVW methods. Horizontal lines denote 95% CI.

We then sought to examine whether there is genetic correlation between RLS and PD that may explain the overrepresentation of these disorders in one another. There was no genetic correlation between RLS and PD (rg=-0.028, se=0.042, *p*=0.507).

## Discussion

Our findings suggest lack of causal relationship between RLS and PD, and lack of genetic correlation. One locus on chromosome 16, including the genes *TOX3* and *CASC16*, is pleiotropic with opposite direction of effect, as SNPs associated with increased risk of RLS are associated with reduced risk of PD, as previously reported.^21^ Therefore, this locus also cannot explain the observed increased frequency of PD in RLS and of RLS in PD.

Although RLS and PD co-occur at a rate higher than expected and share several traits such as dopaminergic treatment response, multiple lines of evidence have shown differences between RLS and PD from a pathophysiological perspective. PD arises from the loss of dopaminergic neurons in the substantia nigra, whereas in RLS there is no loss of dopaminergic cells, no reduced levels of dopamine,^22^ and instead increased presynaptic dopaminergic activity.^23^ The neuronal loss may explain the elevated level of iron (seen as hyperechogenicity in transcranial sonography) and impairment in motor performance in PD versus reduced iron content (hypoechogenicity in transcranial sonography) and normal motor function in idiopathic RLS.^3, 24, 25^.

Our LDSC analyses showed lack of genetic correlation between RLS and PD. Similarly, various genetic studies found no association between known RLS-associated variants and PD in the *BTBD9*^*26,27*^, *MAP2K5*/*SKOR1*^*26,27*^, *MEIS1*^*26,27*^ and *PTPRD*^*26*^ loci. In a study of two Italian families, 10 of 20 RLS patients carried compound heterozygous or single heterozygous *PRKN* variants. It is not clear if these variants are pathogenic, and the clinical symptoms did not differ between RLS patients with and without *PRKN* variants in these 2 families, indicating that their presence was likely random.^28^ In an Asian cohort of 80 PD patients, one patient with a homozygous *PINK1* mutation presented features of RLS, but two other unrelated PD patients with *PINK1* mutations in the same cohort did not show RLS features.^29^ In a study of 258 RLS patients vs 235 healthy controls, the authors reported that the *SNCA* Rep1 allele was associated with reduced risk of RLS.^30^ However, this association was not replicated by the much larger RLS GWAS.^12^ Overall, genetic studies, including the current study, do not support a genetic overlap between RLS and PD.

Our study has some limitations. We could not exclude PD patients with RLS and RLS patients with PD in the datasets used for this analysis, which would have made the results cleaner, since these data was not available. In addition, this study focused on individuals of European ancestry. Studies from multiple ethnicities are required to further study PD, RLS and the association between them. It is possible that rare or structural variants outside of what can be detected with current GWAS technologies are contributing to a shared genetic etiology.

In light of the current and previous findings, it is likely that confounding factors such as treatment, closer neurological follow up and others may have contributed to the observed epidemiological association between RLS and PD. While additional studies are required to identify these potential confounders, the observed association between RLS and PD should not be considered causal on current evidence.

## Data Availability

Available upon request.

## Acknowledgment

We would like to thank all members of the International Parkinson Disease Genomics Consortium (IPDGC). We would like to thank the research participants and employees of 23andMe for making this work possible. For a complete overview of members, acknowledgements and funding, please see http://pdgenetics.org/partners. MAE is funded by the Fonds de Recherche du Québec– Santé (FRQS). GAR holds a Canada Research Chair (Tier 1) in Genetics of the Nervous System and the Wilder Pen field Chair in Neurosciences. ZGO is supported by the Fonds de recherche du Québec–Santé Chercheur-Boursier award and is a Parkinson Canada New Investigator awardee. We thank the International EU-RLS-GENE consortium (https://www.helmholtz-muenchen.de/ing/rls-gene-consortium/members/index.html) for providing RLS GWAS summary statistics. The International EU-RLS-GENE consortium includes data from the COR study which was supported by unrestricted grants to the University of Münster from the German Restless Legs Patient Organisation (RLS Deutsche Restless Legs Vereinigung), the Swiss RLS Patient Association (Schweizerische Restless Legs Selbsthilfegruppe) and a consortium formed by Boeringer Ingelheim Pharma, Mundipharma Research, Neurobiotec, Roche Pharma, UCB (Germany□+□Switzerland) and Vifor Pharma. The clinical material and biospecimens of the Mayo Clinic Florida RLS collection were collected with the assistance of the Mayo Clinic internal funding through the Neuroscience Focused Research Team grant. Genotyping of the International EU-RLS-GENE consortium dataset was supported by DFG grant 218143125 to Prof. Juliane Winkelmann. Participants in the INTERVAL randomised controlled trial were recruited with the active collaboration of NHS Blood and Transplant England (www.nhsbt.nhs.uk), which has supported field work and other elements of the trial. DNA extraction and genotyping were co-funded by the National Institute for Health Research (NIHR), the NIHR BioResource (http://bioresource.nihr.ac.uk/) and the NIHR [Cambridge Biomedical Research Centre at the Cambridge University Hospitals NHS Foundation Trust] [*]. The academic coordinating centre for INTERVAL was supported by core funding from: NIHR Blood and Transplant Research Unit in Donor Health and Genomics (NIHR BTRU-2014-10024), UK Medical Research Council (MR/L003120/1), British Heart Foundation (SP/09/002; RG/13/13/30194; RG/18/13/33946) and the NIHR [Cambridge Biomedical Research Centre at the Cambridge University Hospitals NHS Foundation Trust] [*]. A complete list of the investigators and contributors to the INTERVAL trial is provided in reference.^31^ The academic coordinating centre would like to thank blood donor centre staff and blood donors for participating in the INTERVAL trial and Dr Brendan Burchell (University of Cambridge) for advice on RLS phenotyping. *The views expressed are those of the authors and not necessarily those of the NHS, the NIHR or the Department of Health and Social Care. This work was supported in part by the Intramural Research Program of the National Institute on Aging and National Institute of Neurological Disorders and Stroke (project number Z01-AG000949-02).

## Authors’ Roles

1. Research project: **A**. Conception, **B**. Organization, **C**. Execution
2. Statistical Analysis: **A**. Design, **B**. Execution, **C**. Review and critique
3. Manuscript Preparation: **A**. Writing of the first draft, **B**. Review and critique

MAE: 1A, 1B, 1C, 2A, 2B, 3A

KS: 1C, 2A, 2B, 2C, 3B

EY: 1C, 2B, 2C, 3B

PV: 2B, 2C, 3B

LK: 2A, 3B

SB: 2B, 3B

AJN: 1C, 2B, 2C, 3B

GAR: 1A, 1B, 2C, 3B

ZGO: 1A, 1B, 2A, 2C, 3A, 3B

## Financial disclosure of all authors (for the preceding 12 months)

M.A.E. received doctoral award from Fonds de la recherche en santé du Québec (FRQS). Dr Noyce reports grants from the Barts Charity, Parkinson’s UK, Aligning Science Across Parkinson’s and Michael J Fox Foundation,. Personal fees/honoraria from Britannia, BIAL, AbbVie, Global Kinetics Corporation, Profile, Biogen, Roche and UCB, outside of the submitted work. G.A.R. received grants from the following agencies and organizations; CIHR Foundation Scheme, Genome Quebec, ERANET PerMed/FRSQ, ALS Canada-Brain Canada, ALS Canada, Radala foundation, CQDM, ALS Canada-Brain Canada, Brain Canada. Z.G-O. is supported by the Fonds de recherche du Québec - Santé (FRQS) Chercheurs-boursiers award, and is a Parkinson’s Disease Canada New Investigator awardee. He received consultancy fees from Ono Therapeutics, Handl Therapeutics, Neuron23, Lysosomal Therapeutics Inc., Deerfield, Lighthouse and Idorsia, all unrelated to the current study. All other authors report nothing to disclose.

## Supplementary Tables and Figures

**Supplementary Table 1**. SNPs selected as instrumental variables after clumping.

**Supplementary Figure 1**. Plots showing results from the Mendelian randomization study to evaluate the potential effect of RLS on PD. **A**. Funnel plot indicating estimates of the exposure (RLS) by comparison of results using different MR methods. The effect estimate of the SNP-exposure (RLS) association and the SNP-outcome (PD) associations with standard error bars. Each line corresponds to causal estimates calculated by each MR method. **B**. Funnel plot showing the heterogeneity across the estimates when RLS is exposure. SNPs are represented by dots. IVW and MR Egger computed the average causal effect of all the SNPs. **C**.

**Supplementary Figure 2**. Plots showing results from the Mendelian randomization study to evaluate the potential effect of PD on RLS. **A**. Funnel plot indicating estimates of the exposure (PD) by comparison of results using different MR methods. The effect estimate of the SNP-exposure (PD) association and the SNP-outcome (RLS) associations with standard error bars. Each line corresponds to causal estimates calculated by each MR method. **B**. Funnel plot showing the heterogeneity across the estimates when PD is exposure. SNPs are represented by dots. IVW and MR Egger computed the average causal effect of all the SNPs.

