## Supplementary figures and images for "Lack of Causal Effects or Genetic Correlation between Restless Legs Syndrome and Parkinson’s Disease"

### Supplementary Figure 1

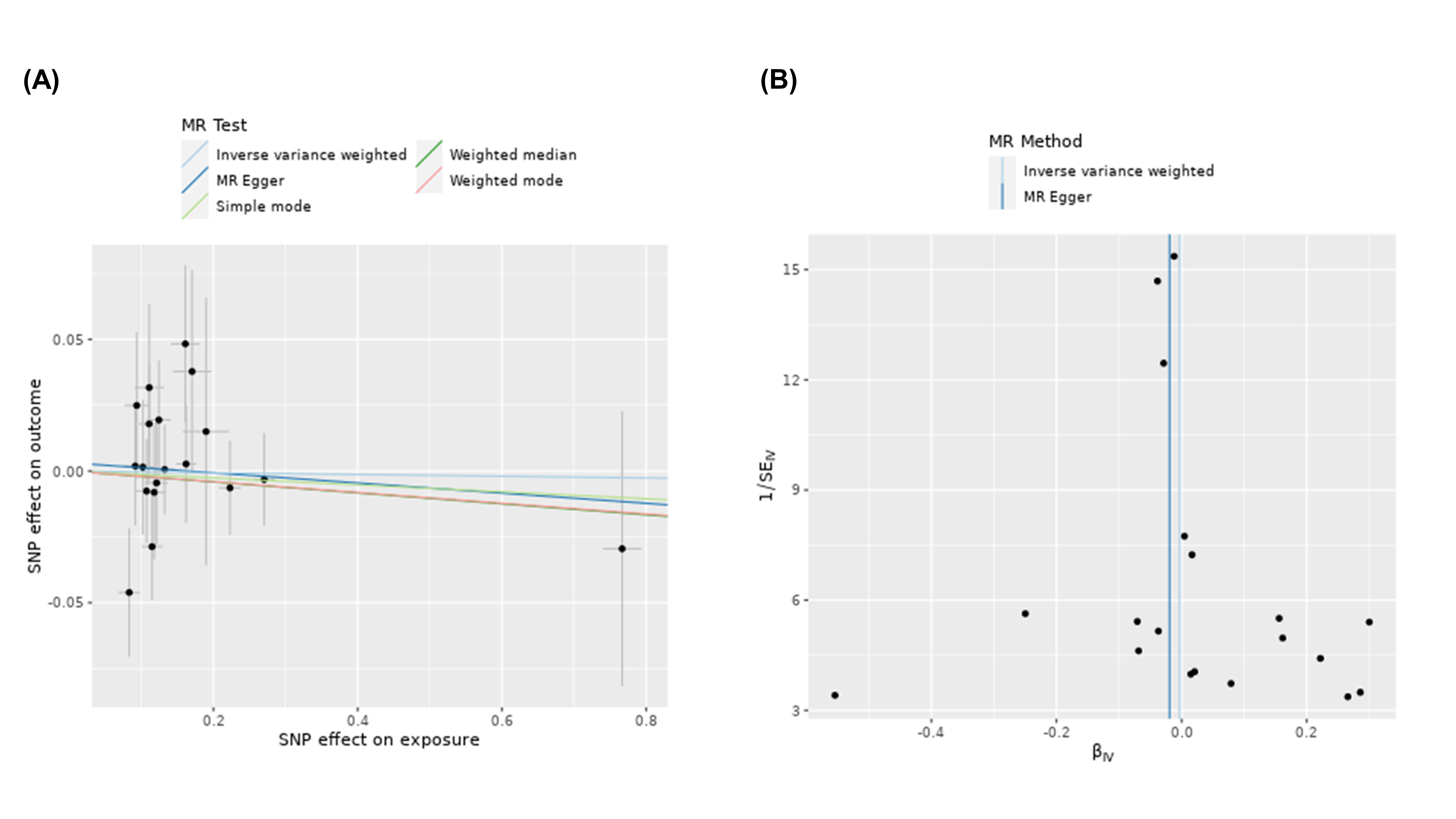

### Supplementary Figure 2

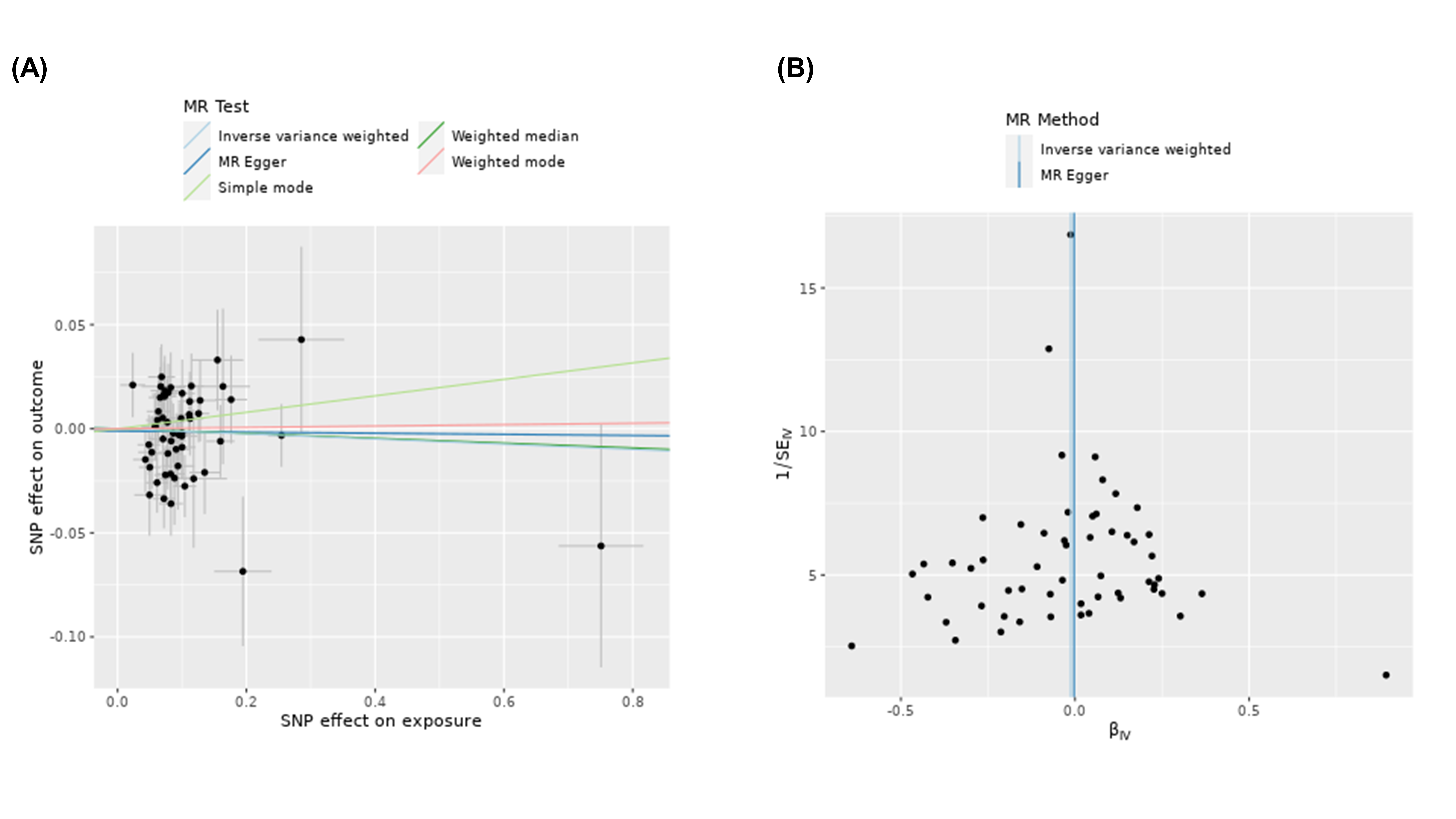
