## Supplementary Table 1 for "Lack of Causal Effects or Genetic Correlation between Restless Legs Syndrome and Parkinson’s Disease"

**Supplementary Table 1**. SNPs selected as instrumental variables for Mendelian randomization analysis.

| **Disease** | **Mapped or nearest gene(s)** | **Clumped SNPs** | **Effect** | **SE** | **Allele Frequency** | **Effect allele** | **Reference allele** | **P value** |
| --- | --- | --- | --- | --- | --- | --- | --- | --- |
| RLS | *AL596327.1, AL499605.1* | rs12046503 | -0.1621 | 0.014 | 0.5887 | T | C | 3.352E-31 |
|  | *LINC01304, AC012445.1* | rs10208712 | 0.1149 | 0.0146 | 0.6404 | A | G | 3.792E-15 |
|  | *MEIS1* | rs113851554 | 0.7668 | 0.0268 | 0.071 | T | G | 1.1E-180 |
|  | *LINC01799* | rs6721312 | 0.0832 | 0.0152 | 0.6927 | A | C | 4.631E-08 |
|  | *AC079112.1, LINC01812* | rs1820989 | -0.1324 | 0.0142 | 0.4685 | A | C | 1.233E-20 |
|  | *CCDC148* | rs80319144 | -0.1244 | 0.0164 | 0.2431 | T | C | 3.193E-14 |
|  | *AC034195.1* | rs1848460 | -0.1178 | 0.0157 | 0.7433 | A | T | 5.39E-14 |
|  | *AC097105.1, RN7SKP212* | rs35987657 | 0.1073 | 0.0148 | 0.666 | A | G | 4.366E-13 |
|  | *LINC02520* | rs74293603 | -0.111 | 0.0202 | 0.1529 | T | C | 3.815E-08 |
|  | *BTBD9* | rs61192259 | 0.2704 | 0.0144 | 0.5945 | A | C | 1.36E-78 |
|  | *AC002069.2* | rs10952927 | -0.161 | 0.0203 | 0.8725 | A | G | 1.861E-15 |
|  | *PTPRD* | rs1836229 | 0.1108 | 0.0139 | 0.5219 | A | G | 1.943E-15 |
|  | *AL139003.1, RPS10P21* | rs340561 | 0.0937 | 0.0171 | 0.2037 | T | G | 3.938E-08 |
|  | *DPH6-DT, LINC02853* | rs996064 | -0.1898 | 0.032 | 0.9446 | A | T | 2.966E-09 |
|  | *AC066615.1, AC073941.2* | rs111652004 | -0.1704 | 0.0264 | 0.096 | T | G | 1.049E-10 |
|  | *MAP2K5* | rs868036 | 0.2229 | 0.0152 | 0.6806 | A | T | 1.108E-48 |
|  | *CASC16* | rs11860769* | 0.2064 | 0.0142 | 0.5838 | A | G | 9.515E-48 |
|  | *LINC02086* | rs12450895 | 0.0916 | 0.0168 | 0.2115 | A | G | 4.884E-08 |
|  | *AC110014.1, LINC01478* | rs12962305 | 0.102 | 0.0159 | 0.2474 | T | C | 1.373E-10 |
|  | *MYT1* | rs365032 | -0.1212 | 0.016 | 0.7298 | A | G | 3.36E-14 |
| PD | *CAB39L* | rs9568188 | 0.0235 | 0.0194 | 0.7444 | T | C | 0.2268 |
|  | *FAM49B* | rs2086641 | -0.0683 | 0.021 | 0.7207 | T | C | 0.001112 |
|  | *BRIP1* | rs61169879 | 0.0671 | 0.0298 | 0.1671 | T | C | 0.02452 |
|  | *LOC100131289* | rs4140646 | 0.0732 | 0.0219 | 0.2221 | A | G | 0.0008346 |
|  | *RPS6KL1* | rs3742785 | 0.0825 | 0.0226 | 0.7863 | A | C | 0.0002597 |
|  | *RETREG3* | rs12951632 | 0.0723 | 0.0189 | 0.7332 | T | C | 0.0001345 |
|  | *GCH1* | rs11158026 | -0.0662 | 0.0181 | 0.3157 | T | C | 0.0002592 |
|  | *MIPOL1* | rs12147950 | -0.0787 | 0.0178 | 0.4357 | T | C | 0.000009459 |
|  | *KCNS3* | rs76116224 | 0.1551 | 0.0403 | 0.9105 | A | T | 0.000119 |
|  | *ITPKB* | rs4653767 | 0.0729 | 0.0186 | 0.7158 | T | C | 0.00008667 |
|  | *FGF20* | rs620513 | -0.1146 | 0.0191 | 0.2675 | T | G | 0.000000002139 |
|  | *SH3GL2* | rs10756907 | -0.1003 | 0.0196 | 0.7598 | A | G | 0.0000003095 |
|  | *RPS12* | rs75859381 | -0.2854 | 0.0668 | 0.97 | T | C | 0.00001954 |
|  | *PAM* | rs26431 | 0.0634 | 0.0183 | 0.7005 | C | G | 0.0005368 |
|  | *CRHR1* | rs117615688 | -0.1636 | 0.0422 | 0.0715 | A | G | 0.0001072 |
|  | *SETD1A* | rs11150601 | 0.112 | 0.0183 | 0.6532 | A | G | 0.0000000009018 |
|  | *VPS13C* | rs2251086 | -0.1282 | 0.0249 | 0.1347 | T | C | 0.000000253 |
|  | *STK39* | rs1474055 | 0.1763 | 0.0248 | 0.1329 | T | C | 0.000000000001144 |
|  | *NOD2* | rs6500328 | 0.0701 | 0.0184 | 0.6022 | A | G | 0.0001353 |
|  | *CHRNB1* | rs12600861 | -0.0619 | 0.0178 | 0.6452 | A | C | 0.6452 |
|  | *TMEM163* | rs57891859 | 0.1112 | 0.019 | 0.7153 | A | G | 0.000000004928 |
|  | *BST1* | rs4698412 | 0.1258 | 0.0168 | 0.553 | A | G | 0.00000000000007049 |
|  | *GPNMB* | rs199351 | 0.0987 | 0.0174 | 0.5907 | A | C | 0.00000001282 |
|  | *IGSF9B* | rs3802920 | 0.1123 | 0.0214 | 0.2123 | T | G | 0.0000001649 |
|  | *RNF141* | rs7938782 | 0.0773 | 0.026 | 0.8729 | A | G | 0.002943 |
|  | *KCNIP3* | rs2042477 | -0.0581 | 0.0215 | 0.2365 | A | T | 0.006891 |
|  | *CLCN3* | rs62333164 | -0.0584 | 0.0183 | 0.3217 | A | G | 0.001435 |
|  | *SNCA* | rs356182 | -0.2545 | 0.021 | 0.6162 | A | G | 9.409E-34 |
|  | *NUCKS1* | rs823118 | 0.0999 | 0.0171 | 0.5748 | T | C | 0.000000004941 |
|  | *LINC00693* | rs6808178 | 0.0864 | 0.0174 | 0.3772 | T | C | 0.0000007199 |
|  | *FAM47E* | rs4101061 | -0.0955 | 0.0186 | 0.7145 | A | G | 0.0000002956 |
|  | *SIPA1L2* | rs10797576 | 0.0998 | 0.0241 | 0.1434 | T | C | 0.00003532 |
|  | *MCCC1* | rs10513789 | 0.1596 | 0.0219 | 0.8174 | T | G | 0.0000000000003185 |
|  | *DNAH17* | rs666463 | 0.0705 | 0.0226 | 0.829 | A | T | 0.001817 |
|  | *KPNA1* | rs55961674 | 0.0832 | 0.0227 | 0.1786 | T | C | 0.0002491 |
|  | *KRTCAP2* | rs35749011 | 0.7508 | 0.0659 | 0.0191 | A | G | 5.022E-30 |
|  | *DYRK1A* | rs2248244 | 0.1001 | 0.0211 | 0.2898 | A | G | 0.000002005 |
|  | *BAG3* | rs72840788 | 0.091 | 0.0206 | 0.2165 | A | G | 0.00001003 |
|  | *VAMP4* | rs11578699 | -0.0781 | 0.0222 | 0.1955 | T | C | 0.0004235 |
|  | *LRRK2* | rs76904798 | 0.1352 | 0.0235 | 0.1486 | T | C | 0.000000009224 |
|  | *IP6K2* | rs12497850 | 0.0485 | 0.0176 | 0.6468 | T | G | 0.005744 |
|  | *CRLS1* | rs77351827 | 0.0937 | 0.0255 | 0.1269 | T | C | 0.0002368 |
|  | *GS1-124K5.11* | rs76949143 | -0.1181 | 0.0517 | 0.0583 | A | T | 0.02232 |
|  | *FYN* | rs997368 | 0.0531 | 0.0211 | 0.8011 | A | G | 0.01192 |
|  | *FAM171A2* | rs850738 | -0.0823 | 0.022 | 0.6041 | A | G | 0.0001887 |
|  | *RIT2* | rs12456492 | -0.1043 | 0.0178 | 0.674 | A | G | 0.000000004885 |
|  | *C5orf24* | rs11950533 | -0.0883 | 0.0283 | 0.1011 | A | C | 0.001813 |
|  | *SCAF11* | rs7134559 | -0.0743 | 0.0172 | 0.4022 | T | C | 0.00001513 |
|  | *UBAP2* | rs6476434 | -0.0431 | 0.0193 | 0.7275 | T | C | 0.02518 |
|  | *SATB1* | rs73038319 | -0.1946 | 0.0446 | 0.9614 | A | C | 0.000013 |
|  | *DLG2* | rs12283611 | -0.05 | 0.017 | 0.4169 | A | C | 0.003228 |
|  | *BIN3* | rs2280104 | 0.0613 | 0.0175 | 0.3636 | T | C | 0.0004527 |
|  | *RIMS1* | rs12528068 | 0.0829 | 0.0186 | 0.2862 | T | C | 0.000008366 |
|  | *MED12L* | rs11707416 | -0.072 | 0.0176 | 0.3697 | A | T | 0.00004533 |
|  | *GBF1* | rs10748818 | -0.0496 | 0.0237 | 0.8524 | A | G | 0.03586 |

*Pleiotropic index SNP. SE, standard error.
